# Changes in essential newborn care practice and maternal and newborn health commodity uptake following implementation of a community-based maternal and newborn care model in South Sudan and Somalia: pre-post study

**DOI:** 10.64898/2026.08.10.26360049

**Authors:** Naoko Kozuki, Mohamed Ahmed Omar, Carolina Cardona, Lual Agok Luka, Grace Kimemia, Geeta Nanda, Asia Mohamed Mohamud, Muna Jama, Chol Peter Deng Yak, Ioanna Wagner Tsoni, Khalisto Baak Wieu, Kur Kur Kur Dut, Lokiri Moses Lowuro, Majok Bol Maduor, Nuoi Yel Dhal, Alom Atak Ayom, Susana Yom Kon Abraham, Abdirisak Dalmar, Teresia Macharia

## Abstract

Given high mortality rates and low access to health facilities, the International Rescue Committee introduced Community-based Maternal and Newborn Care (CBMNC) Programs in rural areas of Somalia and South Sudan. The programs included distribution of evidence-based commodities for maternal or newborn health as well as health counseling during home visits, delivered by low-literate community health workers.

The pre-post study used population-representative cross-sectional surveys among women who delivered in the twelve months preceding the program, conducted before and 18-24 months after the CBMNC program introduction (n=338 baseline, n=340 endline in South Sudan, n=351 baseline, n=302 endline in Somalia). The study employed rigorous statistical methods to adjust for potential confounding factors and strengthen inference regarding changes associated with the program despite the non-experimental study design.

Program enrollment was high (79.1% in South Sudan, 87.7% in Somalia). In South Sudan, Skin-to-skin care, clean cord care, early initiation of breastfeeding, and use of Fansidar were statistically significantly higher among those who received four or more visits, but marginally significantly lower uptake of institutional delivery and SBA. For Somalia, skin-to-skin care showed statistically significant positive change among those who received four or more visits, with early initiation of breastfeeding, no prelacteal feeding, and making four or more facility-based ANC visits demonstrated marginally significant higher uptake.

The positive change in uptake of evidence-based community-based MNH services showed promise in Somalia, but mixed results in South Sudan. This shows promise for change in service uptake even in a relatively short duration of program implementation, but also underscores that community health interventions do not operate in isolation and that parallel investment in facility strengthening and consistent messaging on the complementary roles of community and facility-based care remains essential.

- **What is already known on this topic - *summarise the state of scientific knowledge on this subject before you did your study and why this study needed to be done***

Community-based maternal and newborn care programs have been proven to successfully reduce maternal and newborn mortality and morbidity in development contexts. However, there is limited programming and evidence of a similar type in humanitarian contexts.

- **What this study adds - *summarise what we now know as a result of this study that we did not know before***

Despite a relatively short period of implementation (18-24 months) and delivery of the program by low-literate community health workers, those who received four or more visits from the program showed higher uptake of select maternal and newborn health-related commodities and behaviors. However, in South Sudan, there were marginally significant negative association of program visits with institutional delivery.

- **How this study might affect research, practice or policy - *summarise the implications of this study***

The results show promise for CBMNC programs in humanitarian contexts to increase access and uptake to evidence-based interventions. However, increasing access to community-based services must occur in parallel to strengthening facility-based services to maximize the impact community-based service and counseling delivery can have.

## Background

The impact of fragility and conflict weakens health systems, the consequences of which manifest in high maternal and neonatal mortality. In many fragile and conflict-affected contexts, institutional delivery rates remain low, with countries like Somalia and South Sudan reporting rates of 21–44% [1,2]. To align with commitments for universal health coverage by 2030, more investments need to be made to close this gap in facility-based service access, but the current reality remains that a large proportion of pregnant women cannot reach care.

Community-based maternal and newborn care (CBMNC) programs have demonstrated significant potential for reducing maternal and neonatal mortality. A systematic review [3] highlighted a 20-25% reduction in maternal mortality, maternal morbidity, neonatal mortality, and stillbirths through community-based interventional care packages. Several countries that met their Millennium Development Goals for neonatal mortality reduction [4–7] attribute their successes partially to investments in community health systems.

In Somalia and South Sudan, policy and strategic commitments exist for expanded community-based maternal and newborn health (MNH) care. In Somalia, the *Marwo Caafimaad* program seeks to expand care to women and children at the community level through the establishment of Female Health Workers, and in South Sudan, the national community health policy called the Boma Health Initiative (BHI) includes a mandate to deliver maternal and neonatal interventions. However, as recent political economy analyses on MNH prioritization highlight in both countries [8,9], existing MNH policies have not in practice been followed by political prioritization or financial commitments; the first mention of the *Marwo Caafimaad* program dates back to 2009 and despite a reference to “priority to rural and urban slums,” [10] the program is yet to reach any level of scale in rural contexts and BHI was launched in 2017, but its Safe Motherhood module has rarely been implemented.

Between 2023 and 2025, the International Rescue Committee (IRC) introduced Community-based Maternal and Newborn Care (CBMNC) models, aligned with national community health strategies, in rural areas of Dhusamareb, Somalia, and Northern Bahr El Ghazal, South Sudan. The models aimed to increase uptake of essential newborn care practices, specifically early initiation of breastfeeding, newborn thermal care, and clean cord care, among other behaviors. The programs were not identical but included distribution of evidence-based commodities for MNH as well as health counseling, delivered by low-literate community health workers (CHW). This paper describes the pre-post change at the community level in the uptake of essential newborn care and select MNH-related commodities and behaviors, as well as the change in facility-based service utilization.

## Methods

### Study design

The study was designed as a pre-post study using population-based cross-sectional surveys conducted before and after the introduction of a CBMNC model in South Sudan and Somalia. The analysis employed rigorous statistical methods to adjust for potential confounding factors and strengthen inference regarding changes associated with the program despite the non-experimental study design. The surveys were embedded within a larger implementation research study (presented elsewhere [11]), implemented as part of the EQUAL Research Consortium.

### Program description

The program was implemented from July 2024 to Dec 2025 in South Sudan and Dec 2023 to Dec 2025 in Somalia, a longer implementation period in Somalia. Table 1 describes the CBMNC models, somalias well as the profile of CHWs active in the project, with more details in Appendix File 1.

**Table 1:** Summary of the CBMNC program models and profiles of CHWs in Somalia and South Sudan.

|  | **South Sudan** | **Somalia** |
| --- | --- | --- |
| **Setting** | Four bomas in Aweil East County, Northern Bahr El Ghazal State | Seven rural villages in Dhusamareb district, Galmudug State |
| **Target population** | Pregnant women | Pregnant women |
| **CHW cadre delivering the service** | Boma Health Workers (BHWs), the national CHW cadre operating under the Boma Health Initiative | Female CHWs recruited locally with lower literacy requirements than the national Marwo Caafimaad program |
| **Criteria for CHW selection** | Existing BHWs already operating within the community health system under the Community Health Department as part of the Health Sector Transformation Project | Recruited from within target communities to ensure local acceptability and familiarity with households |
| **Number of CHWs deployed** | 32 BHWs | 34 CHWs |
| **Expected ratio of households served by one CHW** | Initially set as 1:50 per national policy, but household coverage increased with change in policy immediately before rollout (closer to 1:125) | 1:50 |
| **Health messaging provided** | Counselling on anemia prevention in pregnancy (diet and iron/folate adherence); malaria prevention (IPTp-SP and net use); postpartum haemorrhage prevention (misoprostol use); breastfeeding; clean cord care; and newborn thermal care | Counselling on early initiation and exclusive breastfeeding; newborn thermal care (skin-to-skin, delayed bathing, immediate wrapping); clean cord care; nutrition; malaria prevention; hygiene and sanitation; labour signs; immunization |
| **Commodities provided** | Iron–folate tablets; IPTp-SP; misoprostol (for home use after delivery); chlorhexidine gel for cord care | Iron–folate tablets; long-lasting insecticidal nets (LLINs); water purification tablets (Aqua tabs); soap |
| **Supervision mechanism** | Supervisory structure including supervisors and Safe Motherhood Promoters providing mentoring, joint household visits, and reporting support | Supervisory structure including supervisors providing supportive supervision, regular review meetings, and performance feedback |
| **Job aids and tools provided to CHWs** | Pictorial job aids adapted for low literacy; pregnancy cards; BHW registers; simplified service delivery protocols | IEC materials; locally adapted job aids; CHW reporting and data collection tools (including pregnancy mapping tools) |
| **Monitoring data collected by CHWs** | Routine data captured through pregnancy cards, BHW registers, and monthly reporting tools aligned with national systems | Routine data captured through pregnancy mapping tools and CHW reporting systems, including household-level registers and periodic reporting outputs |
| **Complementary community engagement mechanisms** | Structured community engagement activities, including monthly mother-to-mother and father-to-father support groups, quarterly community stakeholder meetings, and radio outreach | Mother-to-Mother and Father-to-Father Support Groups, engagement during existing community health committee meetings and other community stakeholder dialogues, community-based health education sessions |
| **Basic profile of CHWs** |  |  |
| **Gender** | Female: 75%<br>Male: 25% | All female |
| **Age (median; range)** | 35 (28–53) | 28.5 (18–50) |
| **Marital status** | All married | Married 71%<br>Single 15%<br>Divorced/widowed 14% |
| **Educational level** | No formal education: 72%<br>Primary education: 25%<br>Secondary education or above:<br>3% | No formal education: 29%<br>Primary education: 59%<br>Secondary education or above:<br>12% |

While the underlying objectives of the programs were the same, the two programs differed in several ways. First, the CHW cadres differed. In Somalia, *Marwo Caafimaad* was expected to have reached the study location by the time of the study. The *Marwo Caafimaad* program trains women (eligibility requirement of minimum six years of education) over 90 days to deliver maternal, child, and newborn health services [12]. When the program expansion did not proceed, the IRC identified funding to pilot a CBMNC model with a CHW cadre with lower literacy requirements, a profile that aligned more closely with human resource in rural Somalia and acknowledged concerns raised in the 2015 Somali Community Health Strategy [12] that identifying women who meet the educational criteria would be challenging. The initial training for these CHWs was conducted over 20 days, a training timeframe more comparable to CHW programs in other contexts. In South Sudan, Boma Health Workers (BHW), the national CHW cadre already operating in the study area mainly delivering integrated Community Case Management of Childhood Illness services, were recruited. The BHW mandate includes MNH interventions; however, those components of the policy were not being executed systematically or at scale in the country. A three-day refresher training on MNH materials in the existing policy, and an additional ten-day training was conducted for the new model.

Second, the included interventions were different. In both countries, a statistical modeling exercise using a constrained optimization approach to hypothesize on what set of interventions would maximize the number of lives saved was conducted (the results from Somalia are published [13], and the same process was followed for South Sudan). However, there were several interventions in both contexts that were further removed due to policy barriers or feasibility for quick start-up.

Notable difference in profile of the CHWs active in the program include gender (25% women in South Sudan, all women in Somalia) and formal education levels (72% with no formal education in South Sudan, 29% in Somalia).

### Pre-post survey

#### Study population

Women and girls ages 15-49 who reported delivering a baby within the last 12 months were eligible to participate. Informed consent was obtained from all participants. In Somalia, national law recognizes the age of 15 as the age of maturity; girls aged 15 and older were consented as adults. In South Sudan, adolescents aged 15–17 years were eligible to provide independent informed consent in line with ethical approval and national reproductive health policy frameworks, as all adolescent participants were mothers and considered to have the capacity to make informed decisions regarding their care. In both countries, verbal consent was obtained in the local language (Somali in Somalia, Dinka in South Sudan) given low literacy levels.

Women were sampled differently at the two sites, reflecting differences in context and available population data. In Somalia, a pregnancy mapping exercise conducted by CHWs one month prior to each survey identified eligible women, forming the sampling frame. Respondents were then allocated across study villages using probability-proportional-to-size sampling, and simple random sampling was used to select respondents within each village. In South Sudan, where a comparable pregnancy mapping exercise was not feasible, a multistage cluster sampling design was used. Thirty-four village clusters were allocated proportionally across four bomas based on population size, with 10 interviews targeted per cluster. Within each village, households were selected using a modified random walk procedure beginning from a central point, screening every second household. In both contexts, in households with more than one eligible woman, one was randomly selected.

#### Sample size

In Somalia, the sample size of 346 was determined based on detecting a 10 percentage point absolute increase in early initiation of breastfeeding, estimated at 62% according to the 2020 Somalia Health and Demographic Survey, with 80% power and a significance level of 0.05. A total of 351 women were interviewed at baseline and 302 at endline; enough eligible women were not identified at endline, likely due to seasonal out-migration. No women declined participation at baseline and n=1 declined at endline. In South Sudan, the target sample size was 340 women per survey round. The sample was designed to estimate the prevalence of the three primary outcomes (umbilical cord care, thermal care, and early initiation of breastfeeding) with a margin of error of approximately 7.6% at the 95% confidence level, assuming a conservative prevalence of 50% for each outcome and a design effect of 2.0. A total of 338 women were interviewed at baseline, and 340 at endline. The rate of approached individuals who declined to be surveyed was 4.5% at baseline and 1.2% at endline.

#### Data collection

In Somalia, baseline data were collected in October and November 2023 and endline data were collected from November to December 2025. In Somalia, trained female enumerators conducted face-to-face interviews at participants’ homes using the ONA digital data collection platform and Open Data Kit (ODK). The survey tool was developed using indicators from WHO, Demographic and Health Surveys, and other tools used in similar fragile settings, and was translated from English to Somali and back-translated by four bilingual research team members to ensure accuracy. Supervisors conducted spot checks and accompanied enumerators during interviews to monitor adherence to protocols.

In South Sudan, baseline data were collected in March-April 2024 and endline data were collected from October-November 2025. A structured multi-module questionnaire was administered electronically using Computer-Assisted Personal Interviewing (SurveyCTO at baseline, KoboToolbox at endline). The survey was administered in Dinka by South Sudanese data collectors, with concurrent oral translation from English into Dinka to facilitate participant comprehension as written Dinka is not commonly used. Supervisors accompanied enumerators during fieldwork, and daily data quality reviews were conducted to ensure adherence to protocols and data integrity.

#### Variables of interest

Outcome variable definitions (or, as relevant, how they were created) are as follows, and are the same for both countries unless otherwise noted:

*Panel A: Essential newborn care*

- Umbilical cord care

- South Sudan - clean cord care, defined as either did not apply anything to the umbilical cord or applied chlorhexidine AND use a sterilized or new instrument to cut the umbilical cord
- Somalia - Dry cord care, defined as did not apply anything to the umbilical cord

- Thermal care

- Delayed bathing by over 24 hours
- Immediate skin-to-skin contact between newborn and caregiver
- Immediate wrapping of newborn/dried immediately after birth
- All three practices

- Breastfeeding (both countries)

- Early initiation of breastfeeding - Initiation of breastfeeding with the first hour
- Prelacteal feeding – anything other than breastmilk given to the child in first two days

*Panel B: Commodity utilization*

- Iron-folic acid

- Took for 30 or more days
- Took for 90 or more days

- Malaria prevention

- Bednet use during pregnancy
- Intermittent preventive treatment in pregnancy with Sulfadoxine - Pyrimethamine (IPTp-SP)

- Other services

- South Sudan – misoprostol for postpartum hemorrhage

*Panel C: Facility-based care*

- ANC

- Made 1 or more visits
- Made 4 or more visits

- Delivery care

- Institutional delivery
- Skilled birth attendance (SBA)

We did not collect in Somalia whether the instrument to cut the umbilical cord was new or sterilized prior to use, and bednet was only promoted and IPTp-SP was provided in South Sudan, while bednet was provided and IPTp-SP was only promoted in Somalia.

#### Measure of program reach and dose

Exposure to the CBMNC program as the main independent variable of interest was examined in three different ways. First, we measured program coverage using a binary indicator of enrollment in the CBMNC program. Second, we measured contact intensity as the number of prenatal visits received from a CBMNC CHW during pregnancy. This was examined both as a continuous variable and a categorical variable (zero / not enrolled vs. 1-3 visits vs. 4 or more visits). Third, we created a variable we term “dose ratio”; defined as the number of antenatal home visits received divided by the number of months remaining in pregnancy at the time of enrollment, to examine whether a woman received as many visits as she should have when taking into account how early or late she was enrolled in pregnancy. A dose ratio of 1.0 corresponds approximately to one prenatal CHW visit per month remaining in pregnancy at the time of enrollment.

These measures were only collected among women interviewed at endline. For analyses that pooled baseline and endline observations, we assigned a value of zero to all three measures for baseline respondents.

#### Demographic, socioeconomic, and reproductive health covariates

Demographic variables included maternal age at time of survey (15-29 vs. 30-49 years), age at first marriage (15-17 vs. ≥18 years), and total number of pregnancies at time of survey. Socioeconomic variables included maternal education (no schooling/Quranic school vs. primary education or higher), household wealth (lower vs. higher, derived using a principal components analysis–based asset index; Appendix File 2), travel time to the nearest health facility (<30 minutes, 30–<60 minutes, 1–2 hours, and ≥2 hours), and primary mode of transportation to a health facility (walking vs. motorized or bicycle transport). We also adjusted for pregnancy-related and reproductive health characteristics, including decision-making autonomy for medical care (respondent alone vs. husband/partner/other), fertility intentions for the index pregnancy, awareness of birth spacing methods, and previous use of family planning methods. To account for geographic access to services, models additionally included the natural logarithm of the minimum GPS-derived distance from a woman’s household to any health facility.

### Statistical analysis

#### Descriptive analysis and unadjusted pre-post comparison

We first summarized the demographic, socioeconomic, reproductive health, and healthcare access characteristics of women included in the analytical sample to describe the composition of the study population at baseline and endline, comparing the unadjusted prevalence of the characteristics. Then, we estimated the prevalence of all outcomes included in the evaluation separately at baseline and endline, encompassing three domains: (i) Essential Newborn Care practices, including thermal care, clean cord care, and breastfeeding practices; (ii) MNH commodity utilization, including iron-folic acid supplementation, bednet use, and IPTp-SP uptake; and (iii) facility-based maternal health service utilization, including receipt of at least one antenatal care (ANC) visit, receipt of at least four ANC visits, SBA, and institutional delivery. We compared the unadjusted prevalence of these outcomes across baseline and endline survey round.

We described exposure to the CBMNC program among endline respondents, including enrollment in the program and the number of prenatal CBMNC visits received during pregnancy. To assess differences in participant characteristics across levels of program exposure, we tabulated respondent characteristics by three categories of antenatal CBMNC visits received (none, 1–3 visits, and 4 or more visits).

#### Adjusted regression models with CBMNC exposure as main independent variable

We examined changes in MNH outcomes between baseline and endline using separate modified Poisson regression models with a log link for each binary outcome. Huber–White robust standard errors were used to account for variance misspecification arising from the application of a Poisson model to binary data, yielding valid estimates of risk ratios (RRs) and 95% confidence intervals. We estimated models with standard errors clustered at the village level and compared them with models that included village fixed effects to account for time-invariant differences across villages and robust standard errors. The specification including village fixed effects had a lower Akaike Information Criterion and was therefore selected for the primary analyses.

To quantify changes in MNH outcomes between baseline and endline, we first estimated the following model:

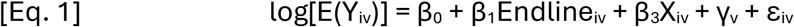

where Y_iv_ represents a binary maternal or newborn health outcome for woman *i* residing in village *v*; Endline_iv_ is an indicator variable equal to 1 for observations from the endline survey and 0 for observations from the baseline survey; X_iv_ is a vector of demographic, socioeconomic, and reproductive health covariates; and γ_v_ represents village fixed effects. Exponentiated coefficients are interpreted as adjusted RRs. The exponentiated coefficient of β_1_ estimates the adjusted RR comparing the prevalence of the outcome at endline with its prevalence at baseline, thereby quantifying the extent of change in the outcome between survey rounds after adjustment for observed covariates.

To assess whether exposure to the CBMNC program was associated with MNH outcomes among women observed at endline, we estimated the following model:

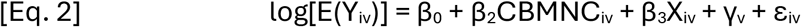

where CBMNC_iv_ is a categorical variable indicating the number of prenatal CBMNC visits received by woman *i* (none, 1–3 visits, or 4 or more visits). In this model, the exponentiated coefficients associated with the CBMNC categories (β_2_) estimate the adjusted RRs comparing the prevalence of the outcome among women receiving 1-3 visits or 4 or more visits with that among women receiving no CBMNC visits, after adjustment for observed covariates and village fixed effects. This equation only utilizes data from the endline survey, where all surveyed respondents had the chance to be enrolled in the program.

To estimate the association between CBMNC exposure and MNH outcomes while accounting for underlying changes between baseline and endline, we estimated the following model:

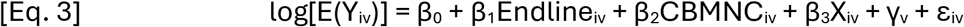

In this model, the exponentiated coefficient of β_1_ estimates the adjusted RR comparing the prevalence of the outcome at endline with its prevalence at baseline after accounting for CBMNC exposure and other covariates. The exponentiated coefficients associated with the CBMNC categories (β_2_) estimate the adjusted RRs comparing the prevalence of the outcome among women receiving 1–3 visits or 4 or more visits with that among women receiving no CBMNC visits, after adjustment for survey round and other covariates. These estimates can be interpreted as the association between CBMNC exposure and the outcome, net of temporal changes between baseline and endline.

The primary analyses reported in this manuscript are based on Equation 3. Regression results from Equations 1 and 2 are presented in Appendix File 3 and are included to characterize changes in outcomes between baseline and endline and associations between CBMNC exposure and outcomes among endline respondents. Exponentiated coefficients from all models are presented as adjusted RRs.

All models adjusted for maternal age, age at marriage, parity, maternal education, travel time to the nearest health facility, mode of transportation, healthcare decision-making, fertility intentions for the last birth, awareness of birth spacing methods, prior use of family planning, and the distance to the nearest health facility. Models estimating Essential Newborn Care outcomes, MNH Commodity Utilization outcomes, SBA, and institutional delivery additionally adjusted for receipt of at least one facility-based antenatal care visit during pregnancy. Collinearity testing was conducted to avoid inclusion of collinear variables.

All Appendix Tables and Figures are available in Appendix File 4.

## Research involvement

Researchers from the IRC played a major part in the research methodology and quality assurance of the data collection, but did not collect any data directly and were not directly involved in the service delivery. Data analysis was conducted by a researcher outside of the IRC. Research subjects or members of the public were not involved in the design, or conduct, or reporting, or dissemination plans of the research.

## Ethical approval

The studies were approved by Somali Research and Development Institute IRB and Somalia Ministry of Health for the Somalia study and by the South Sudan Ministry of Health for the South Sudan study, the IRC IRB for both studies.

## Results

351 women at baseline and 302 women at endline consented to the survey in Somalia and 338 women at baseline and 340 women at endline in South Sudan. The characteristics of the surveyed women are available in Table 2. Notably, characteristics like age at first marriage, travel time to a health facility, and several variables around family planning were statistically significantly different between baseline and endline in South Sudan and several education and wealth-related variables, travel time, and several variables around family planning in Somalia. Distribution of household wealth index scores is available in Appendix Figure 1.

**Table 2:** Characteristics of surveyed women, stratified by baseline/endline and by location.

| Variables | South Sudan |  |  |  |  | Somalia |  |  |  |  |
| --- | --- | --- | --- | --- | --- | --- | --- | --- | --- | --- |
|  | Baseline |  | Endline |  | p-value | Baseline |  | Endline |  | p-value |
|  | Prop. | Obs. | Prop. | Obs. |  | Prop. | Obs. | Prop. | Obs. |  |
| Mother's age (years) |  |  |  |  |  |  |  |  |  |  |
| Less than 29 | 67.2 | 227 | 66.2 | 225 | 0.786 | 59.8 | 210 | 65.9 | 199 | 0.110 |
| 30 and above | 32.8 | 111 | 33.8 | 115 |  | 40.2 | 141 | 34.1 | 103 |  |
| Age at 1st marriage |  |  |  |  |  |  |  |  |  |  |
| 18 and above | 37.5 | 126 | 49.4 | 168 | **0.002** | 39.1 | 134 | 44.0 | 133 | 0.201 |
| Less than 18 | 62.5 | 210 | 50.6 | 172 |  | 60.9 | 209 | 56.0 | 169 |  |
| Marital status |  |  |  |  |  |  |  |  |  |  |
| Married | 97.6 | 322 | 97.3 | 319 | 0.796 | 94.3 | 331 | 95.4 | 288 | 0.542 |
| Divorced/Widowed | 2.4 | 8 | 2.7 | 9 |  | 5.7 | 20 | 4.6 | 14 |  |
| Father's age (years) |  |  |  |  |  |  |  |  |  |  |
| Less than 29 | 13.5 | 42 | 12.9 | 43 | 0.812 | 20.3 | 48 | 26.9 | 73 | **0.082** |
| 30 and above | 86.5 | 268 | 87.1 | 290 |  | 79.7 | 188 | 73.1 | 198 |  |
| Mother's education |  |  |  |  |  |  |  |  |  |  |
| No schooling/Quranic school | 70.7 | 239 | 68.5 | 233 | 0.537 | 81.5 | 286 | 71.9 | 217 | **0.004** |
| Primary/secondary/higher | 29.3 | 99 | 31.5 | 107 |  | 18.5 | 65 | 28.1 | 85 |  |
| Father's education |  |  |  |  |  |  |  |  |  |  |
| No schooling/Quranic school | 47.0 | 158 | 52.5 | 177 | 0.154 | 74.8 | 208 | 63.9 | 175 | **0.005** |
| Primary/secondary/higher | 53.0 | 178 | 47.5 | 160 |  | 25.2 | 70 | 36.1 | 99 |  |
| Wealth |  |  |  |  |  |  |  |  |  |  |
| Lowest | 50.0 | 169 | 50.0 | 170 | 1.000 | 50.1 | 176 | 50.0 | 151 | 0.971 |
| Highest | 50.0 | 169 | 50.0 | 170 |  | 49.9 | 175 | 50.0 | 151 |  |
| Travel time to health facility |  |  |  |  |  |  |  |  |  |  |
| <30 minutes | 4.4 | 15 | 10.9 | 37 | **<0.001** | 19.3 | 64 | 24.2 | 72 | **<0.001** |
| 30-<60 minutes | 6.5 | 22 | 20.0 | 68 |  | 23.3 | 77 | 26.3 | 78 |  |
| 1-2 hours | 34.3 | 116 | 42.6 | 145 |  | 40.8 | 135 | 43.4 | 129 |  |
| 2+ hours | 54.7 | 185 | 26.5 | 90 |  | 16.6 | 55 | 6.1 | 18 |  |
| Mode of transportation to a health facility |  |  |  |  |  |  |  |  |  |  |
| Car/Truck/Motorcycle/Bicycle | 3.8 | 13 | 4.7 | 16 | 0.575 | 78.0 | 270 | 75.7 | 227 | 0.476 |
| Walking | 96.2 | 325 | 95.3 | 323 |  | 22.0 | 76 | 24.3 | 73 |  |
| Decision on medical care |  |  |  |  |  |  |  |  |  |  |
| Respondent | 21.6 | 73 | 13.5 | 46 | **0.006** | 39.9 | 140 | 24.5 | 80 | **<0.001** |
| Husband/partner/Other | 78.4 | 265 | 86.5 | 294 |  | 60.1 | 211 | 75.5 | 247 |  |
| Fertility intentions of last pregnancy |  |  |  |  |  |  |  |  |  |  |
| No | 20.1 | 68 | 9.4 | 32 | **<0.001** | 17.3 | 59 | 9.7 | 29 | **0.006** |
| Yes | 79.9 | 270 | 90.6 | 308 |  | 82.7 | 283 | 90.3 | 270 |  |
| Awareness of birth spacing methods |  |  |  |  |  |  |  |  |  |  |
| No | 62.1 | 210 | 85.9 | 292 | **<0.001** | 74.9 | 263 | 53.6 | 162 | **<0.001** |
| Yes | 37.9 | 128 | 14.1 | 48 |  | 25.1 | 88 | 46.4 | 140 |  |
| Previous use of FP methods |  |  |  |  |  |  |  |  |  |  |
|  | Baseline |  | Endline |  | p-value | Baseline |  | Endline |  | p-value |
|  | Prop. | Obs. | Prop. | Obs. |  | Prop. | Obs. | Prop. | Obs. |  |
| No | 77.8 | 263 | 85.3 | 290 | **0.012** | 83.8 | 294 | 71.5 | 216 | **<0.001** |
| Yes | 22.2 | 75 | 14.7 | 50 |  | 16.2 | 57 | 28.5 | 86 |  |
| Number of total pregnancies | 4.1 |  | 3.9 |  | 0.135 | 5.9 |  | 5.6 |  | 0.147 |

Notes: P-values for categorical variables were based on Pearson's $\chi^2$ tests comparing baseline and endline distributions within each country. P-values for continuous variables, where applicable, were based on two-sample t-tests. The denominators do not necessarily equal the same across all variables due to "don't know" responses that were excluded from this table.

Table 3 represents the unadjusted temporal change between baseline and endline, reporting crude prevalence at baseline and endline and an unadjusted risk ratio of change over time. In South Sudan, all outcomes except early initiation of breastfeeding, IFA for 30 or more days, four or more facility-based ANC visits, and SBA statistically significantly improved from baseline to endline. Of note, chlorhexidine which was a newly introduced drug in the community had an uptake of 90%. Practicing all three thermal care actions had the highest unadjusted change, going from 5% to 46%. All indicators statistically significantly improved in Somalia, with the largest relative change seen in IFA utilization (30-day use of IFA from 10 % to 68%, 90-day use from 1% to 15%) and thermal care (all three behaviors, from 15% to 43%). The same data as Table 2 and 3 stratified by categorical exposure to the CBMNC program (0 vs. 1-3 vs. 4 or more home visits) is available as Appendix Tables 3 and 4. Appendix Table 1 describes the reported items applied to the cord and prelacteal foods given. Data on exposure to counseling on breastfeeding, umbilical cord care, and thermal care are available in Appendix Table 2, with all three statistically significantly increasing.

**Table 3:**
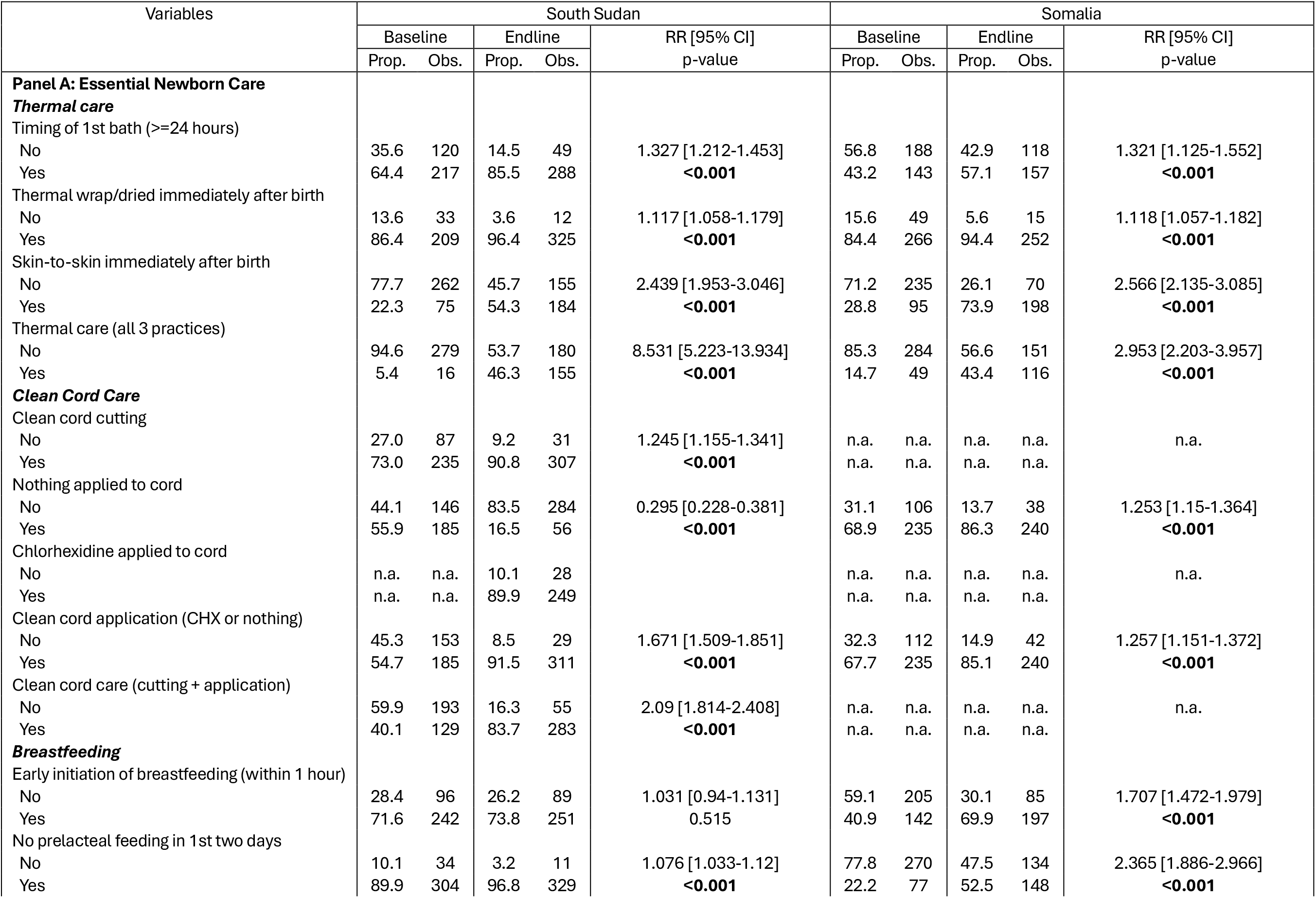
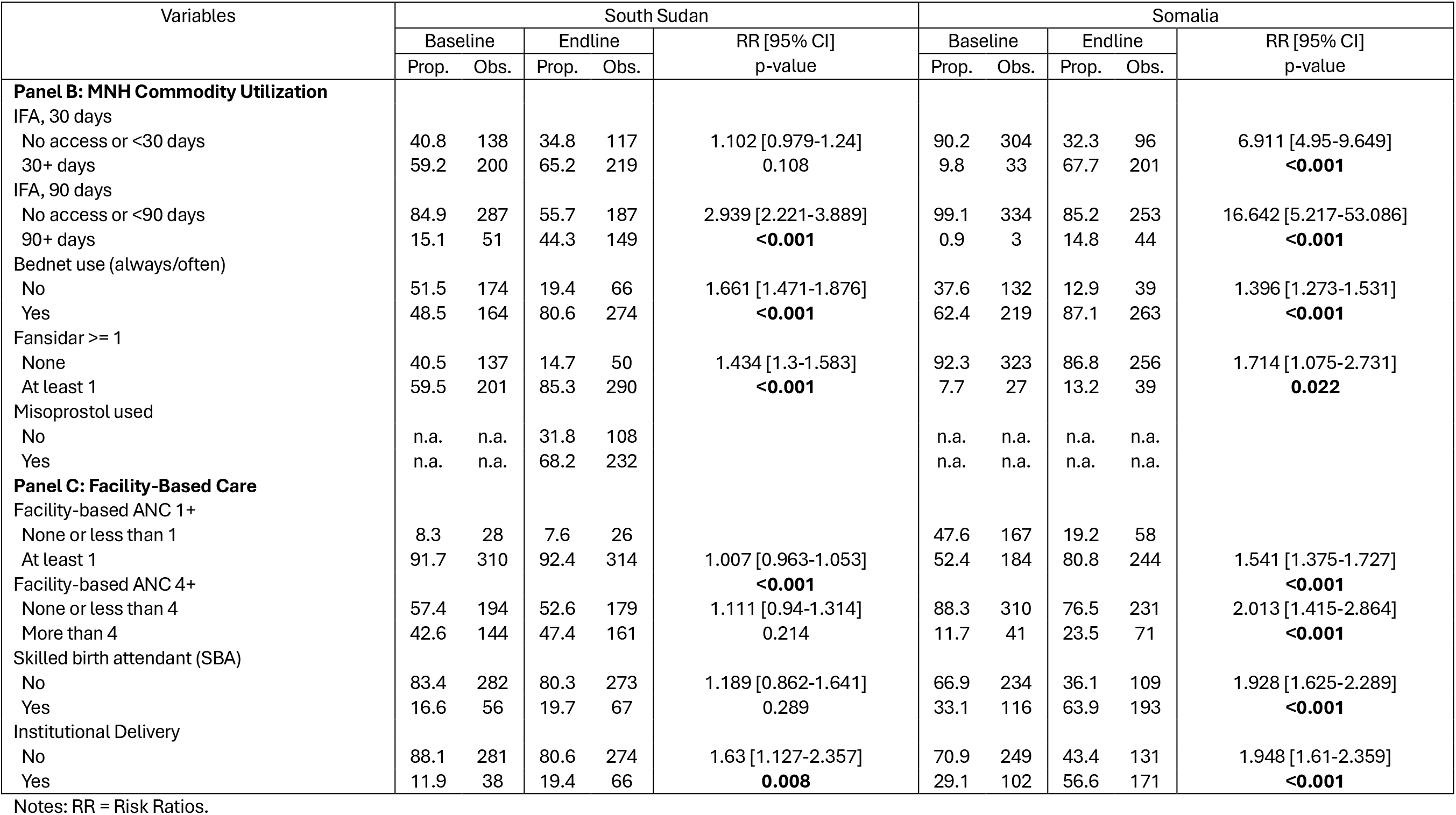
Temporal change in essential newborn care practices, MNH commodity utilization, and facility-based maternal care between baseline and endline in South Sudan and Somalia.

| Variables | South Sudan |  |  |  |  | Somalia |  |  |  |  |
| --- | --- | --- | --- | --- | --- | --- | --- | --- | --- | --- |
|  | Baseline |  | Endline |  | RR [95% CI]<br>p-value | Baseline |  | Endline |  | RR [95% CI]<br>p-value |
|  | Prop. | Obs. | Prop. | Obs. |  | Prop. | Obs. | Prop. | Obs. |  |
| **Panel A: Essential Newborn Care** |  |  |  |  |  |  |  |  |  |  |
| ***Thermal care*** |  |  |  |  |  |  |  |  |  |  |
| Timing of 1st bath (>=24 hours) |  |  |  |  |  |  |  |  |  |  |
| No | 35.6 | 120 | 14.5 | 49 | 1.327 [1.212-1.453] | 56.8 | 188 | 42.9 | 118 | 1.321 [1.125-1.552] |
| Yes | 64.4 | 217 | 85.5 | 288 | <0.001 | 43.2 | 143 | 57.1 | 157 | <0.001 |
| Thermal wrap/dried immediately after birth |  |  |  |  |  |  |  |  |  |  |
| No | 13.6 | 33 | 3.6 | 12 | 1.117 [1.058-1.179] | 15.6 | 49 | 5.6 | 15 | 1.118 [1.057-1.182] |
| Yes | 86.4 | 209 | 96.4 | 325 | <0.001 | 84.4 | 266 | 94.4 | 252 | <0.001 |
| Skin-to-skin immediately after birth |  |  |  |  |  |  |  |  |  |  |
| No | 77.7 | 262 | 45.7 | 155 | 2.439 [1.953-3.046] | 71.2 | 235 | 26.1 | 70 | 2.566 [2.135-3.085] |
| Yes | 22.3 | 75 | 54.3 | 184 | <0.001 | 28.8 | 95 | 73.9 | 198 | <0.001 |
| Thermal care (all 3 practices) |  |  |  |  |  |  |  |  |  |  |
| No | 94.6 | 279 | 53.7 | 180 | 8.531 [5.223-13.934] | 85.3 | 284 | 56.6 | 151 | 2.953 [2.203-3.957] |
| Yes | 5.4 | 16 | 46.3 | 155 | <0.001 | 14.7 | 49 | 43.4 | 116 | <0.001 |
| ***Clean Cord Care*** |  |  |  |  |  |  |  |  |  |  |
| Clean cord cutting |  |  |  |  |  |  |  |  |  |  |
| No | 27.0 | 87 | 9.2 | 31 | 1.245 [1.155-1.341] | n.a. | n.a. | n.a. | n.a. | n.a. |
| Yes | 73.0 | 235 | 90.8 | 307 | <0.001 | n.a. | n.a. | n.a. | n.a. |  |
| Nothing applied to cord |  |  |  |  |  |  |  |  |  |  |
| No | 44.1 | 146 | 83.5 | 284 | 0.295 [0.228-0.381] | 31.1 | 106 | 13.7 | 38 | 1.253 [1.15-1.364] |
| Yes | 55.9 | 185 | 16.5 | 56 | <0.001 | 68.9 | 235 | 86.3 | 240 | <0.001 |
| Chlorhexidine applied to cord |  |  |  |  |  |  |  |  |  |  |
| No | n.a. | n.a. | 10.1 | 28 |  | n.a. | n.a. | n.a. | n.a. | n.a. |
| Yes | n.a. | n.a. | 89.9 | 249 |  | n.a. | n.a. | n.a. | n.a. |  |
| Clean cord application (CHX or nothing) |  |  |  |  |  |  |  |  |  |  |
| No | 45.3 | 153 | 8.5 | 29 | 1.671 [1.509-1.851] | 32.3 | 112 | 14.9 | 42 | 1.257 [1.151-1.372] |
| Yes | 54.7 | 185 | 91.5 | 311 | <0.001 | 67.7 | 235 | 85.1 | 240 | <0.001 |
| Clean cord care (cutting + application) |  |  |  |  |  |  |  |  |  |  |
| No | 59.9 | 193 | 16.3 | 55 | 2.09 [1.814-2.408] | n.a. | n.a. | n.a. | n.a. | n.a. |
| Yes | 40.1 | 129 | 83.7 | 283 | <0.001 | n.a. | n.a. | n.a. | n.a. |  |
| ***Breastfeeding*** |  |  |  |  |  |  |  |  |  |  |
| Early initiation of breastfeeding (within 1 hour) |  |  |  |  |  |  |  |  |  |  |
| No | 28.4 | 96 | 26.2 | 89 | 1.031 [0.94-1.131] | 59.1 | 205 | 30.1 | 85 | 1.707 [1.472-1.979] |
| Yes | 71.6 | 242 | 73.8 | 251 | 0.515 | 40.9 | 142 | 69.9 | 197 | <0.001 |
| No prelacteal feeding in 1st two days |  |  |  |  |  |  |  |  |  |  |
| No | 10.1 | 34 | 3.2 | 11 | 1.076 [1.033-1.12] | 77.8 | 270 | 47.5 | 134 | 2.365 [1.886-2.966] |
| Yes | 89.9 | 304 | 96.8 | 329 | <0.001 | 22.2 | 77 | 52.5 | 148 | <0.001 |
|  | Baseline |  | Endline |  | RR [95% CI]<br>p-value | Baseline |  | Endline |  | RR [95% CI]<br>p-value |
|  | Prop. | Obs. | Prop. | Obs. |  | Prop. | Obs. | Prop. | Obs. |  |
| **Panel B: MNH Commodity Utilization** |  |  |  |  |  |  |  |  |  |  |
| IFA, 30 days |  |  |  |  |  |  |  |  |  |  |
| No access or <30 days | 40.8 | 138 | 34.8 | 117 | 1.102 [0.979-1.24] | 90.2 | 304 | 32.3 | 96 | 6.911 [4.95-9.649] |
| 30+ days | 59.2 | 200 | 65.2 | 219 | 0.108 | 9.8 | 33 | 67.7 | 201 | <0.001 |
| IFA, 90 days |  |  |  |  |  |  |  |  |  |  |
| No access or <90 days | 84.9 | 287 | 55.7 | 187 | 2.939 [2.221-3.889] | 99.1 | 334 | 85.2 | 253 | 16.642 [5.217-53.086] |
| 90+ days | 15.1 | 51 | 44.3 | 149 | <0.001 | 0.9 | 3 | 14.8 | 44 | <0.001 |
| Bednet use (always/often) |  |  |  |  |  |  |  |  |  |  |
| No | 51.5 | 174 | 19.4 | 66 | 1.661 [1.471-1.876] | 37.6 | 132 | 12.9 | 39 | 1.396 [1.273-1.531] |
| Yes | 48.5 | 164 | 80.6 | 274 | <0.001 | 62.4 | 219 | 87.1 | 263 | <0.001 |
| Fansidar >= 1 |  |  |  |  |  |  |  |  |  |  |
| None | 40.5 | 137 | 14.7 | 50 | 1.434 [1.3-1.583] | 92.3 | 323 | 86.8 | 256 | 1.714 [1.075-2.731] |
| At least 1 | 59.5 | 201 | 85.3 | 290 | <0.001 | 7.7 | 27 | 13.2 | 39 | 0.022 |
| Misoprostol used |  |  |  |  |  |  |  |  |  |  |
| No | n.a. | n.a. | 31.8 | 108 |  | n.a. | n.a. | n.a. | n.a. |  |
| Yes | n.a. | n.a. | 68.2 | 232 |  | n.a. | n.a. | n.a. | n.a. |  |
| **Panel C: Facility-Based Care** |  |  |  |  |  |  |  |  |  |  |
| Facility-based ANC 1+ |  |  |  |  |  |  |  |  |  |  |
| None or less than 1 | 8.3 | 28 | 7.6 | 26 |  | 47.6 | 167 | 19.2 | 58 |  |
| At least 1 | 91.7 | 310 | 92.4 | 314 | 1.007 [0.963-1.053] | 52.4 | 184 | 80.8 | 244 | 1.541 [1.375-1.727] |
| Facility-based ANC 4+ |  |  |  |  | <0.001 |  |  |  |  | <0.001 |
| None or less than 4 | 57.4 | 194 | 52.6 | 179 | 1.111 [0.94-1.314] | 88.3 | 310 | 76.5 | 231 | 2.013 [1.415-2.864] |
| More than 4 | 42.6 | 144 | 47.4 | 161 | 0.214 | 11.7 | 41 | 23.5 | 71 | <0.001 |
| Skilled birth attendant (SBA) |  |  |  |  |  |  |  |  |  |  |
| No | 83.4 | 282 | 80.3 | 273 | 1.189 [0.862-1.641] | 66.9 | 234 | 36.1 | 109 | 1.928 [1.625-2.289] |
| Yes | 16.6 | 56 | 19.7 | 67 | 0.289 | 33.1 | 116 | 63.9 | 193 | <0.001 |
| Institutional Delivery |  |  |  |  |  |  |  |  |  |  |
| No | 88.1 | 281 | 80.6 | 274 | 1.63 [1.127-2.357] | 70.9 | 249 | 43.4 | 131 | 1.948 [1.61-2.359] |
| Yes | 11.9 | 38 | 19.4 | 66 | 0.008 | 29.1 | 102 | 56.6 | 171 | <0.001 |

Notes: RR = Risk Ratios.

Appendix Figure 2 summarizes exposure to the CBMNC program. Enrollment was high in both countries, reaching 79.1% in South Sudan and 87.7% in Somalia. Program intensity was greater in Somalia, where 71.6% of women received four or more antenatal CBMNC visits compared with 42.1% in South Sudan, while a smaller proportion of women received no visits (11.9% vs. 21.5%). Consistent with these findings, the median antenatal CBMNC dose ratio was substantially higher in Somalia (0.94) than in South Sudan (0.50), indicating that women in Somalia received nearly one prenatal CBMNC visit per month of pregnancy after enrollment, compared with approximately one visit every two months in South Sudan.

Figure 1 presents the adjusted regression results, comparing individuals who received zero visits from the CBMNC program (reference) versus 1-3 visits versus four or more visits antenatally for South Sudan and Somalia (Equation 3). Table 4 describes the regression results for uptake of IFA and malaria prevention. In South Sudan, skin-to-skin care, clean cord care, early initiation of breastfeeding, and use of IPTp-SP all experienced a statistically significant change. Notably, institutional delivery and SBA were both marginally significantly (p<0.10) lower by 41% (aRR 0.59, 95% CI: 0.32 – 1.08) among those receiving 1-3 visits and 42% (aRR 0.57, 95% CI: 0.32 - 1.03) respectively among those who received four or more visits. However, it is worth noting the minor difference in the absolute value of both outcomes in the unadjusted analysis, comparing those with no versus 1-3 versus 4 or more visits (institutional delivery: 23.3% vs. 18.5% vs. 18.2%, SBA: 24.7% vs. 18.5% vs. 18.2%).

**Figure 1:**
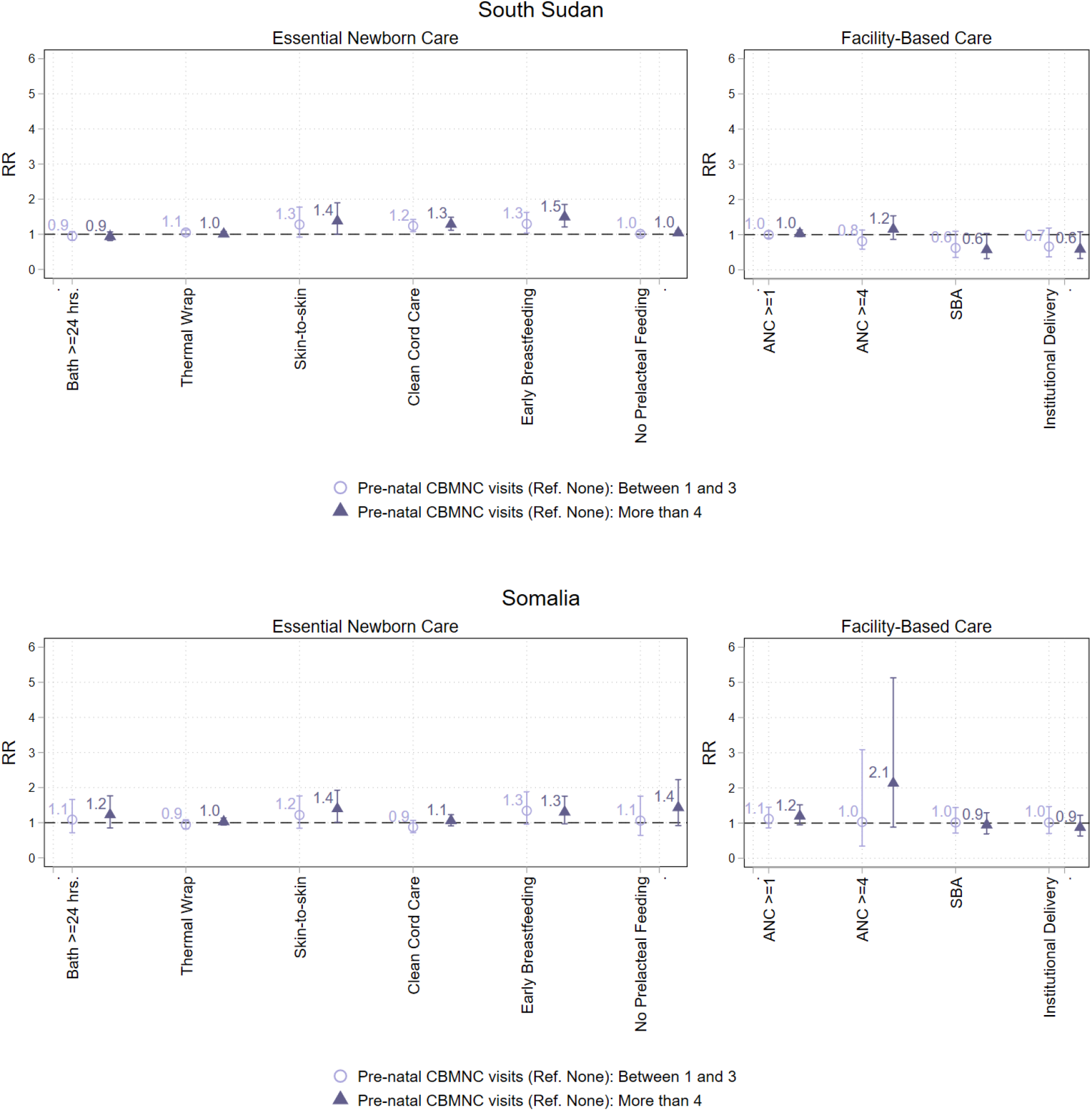
Association between antenatal CBMNC visits received (0 as reference vs. 1-3 visits vs. 4 or more visits) and key outcome indicators for South Sudan (top) and Somalia (bottom)

**Table 4:** Adjusted risk ratios of CBMNC visits received and outcomes related to IFA use and malaria prevention behaviors.

| Outcome | Pre-natal<br>CBMNC visits<br>(Ref. None) | South Sudan |  | Somalia |  |
| --- | --- | --- | --- | --- | --- |
|  |  | RR [95% CI] | Obs. | RR [95% CI] | Obs. |
| IFA 30 days | 1-3 visits | 1.306<br>[1.038 - 1.642] | 670 | 2.027<br>[1.237 - 3.322] | 596 |
|  | 4+ | 1.414<br>[1.139 - 1.756] |  | 2.339<br>[1.484 - 3.686] |  |
| IFA 90 days | 1-3 visits | 1.600<br>[1.056 - 2.424] | 670 | N/A* | 596 |
|  | 4+ | 2.148<br>[1.459 - 3.162] |  | N/A* |  |
| Fansidar use** | 1-3 visits | 1.680<br>[1.356 - 2.082] | 674 | 1.709<br>[0.382 - 7.642] | 605 |
|  | 4+ | 1.706<br>[1.380 - 2.109] |  | 2.112<br>[0.525 - 8.499] |  |
| LLITN use*** | 1-3 visits | 1.040<br>[0.887 - 1.221] | 674 | 1.187<br>[0.949 - 1.484] | 613 |
|  | 4+ | 1.052<br>[0.899 - 1.231] |  | 1.177<br>[0.962 - 1.439] |  |

Notes: Risk ratios (RRs) and 95% confidence intervals are derived from the CBMNC exposure coefficients ( $\beta_2$ ) estimated in Equation 3 using modified Poisson regression models adjusted for survey round, demographic, socioeconomic, reproductive health, and village-level characteristics. Reference group: women with no prenatal CBMNC visits.
\*Only n=3 reported using IFA for 90+ days in Somalia at baseline, making the regression results unstable, and therefore were not included here.
\*\*IPTp-SP was offered as part of the package in South Sudan, but not in Somalia.
\*\*\*LLITN was offered as part of the package in Somalia, but not in South Sudan.

For Somalia, skin-to-skin care showed statistically significant positive change at the p<0.05 level, and early initiation of breastfeeding, no prelacteal feeding, and making four or more facility-based ANC visits at the p<0.10 level. Somalia did not show a statistically significant change, negative or positive, in SBA or institutional delivery.

There was statistically significant higher IFA use for 30 days in both countries, and in IFA 90 days and IPTp-SP use in South Sudan (Table 4).

## Discussion

In a context where access to critical life-saving maternal and newborn care is severely limited, new pilot programs for community-based MNH service delivery in rural contexts of South Sudan and Somalia showed high coverage. The change in uptake of evidence-based Essential Newborn Care behaviors and other MNH commodities and services before and after the implementation of the CBMNC program showed promise in increasing key newborn care behaviors in Somalia, but mixed results in South Sudan, where program engagement was associated with higher uptake of certain newborn care behaviors but lower uptake of institutional delivery. Reflections on how to operationalize such community-based programs will be available in separate implementation research results [11].

### Lower association with institutional delivery in South Sudan

Our analysis found a marginally significant inverse association between the number of CHW visits received and institutional delivery in South Sudan; however, the absolute difference in institutional delivery rates was modest. Much of the literature suggests that facility-based antenatal care is positively associated with facility delivery [14–16], highlighting a potentially differential pathway between facility-based antenatal care versus antenatal home visits and the outcome of institutional delivery. A well-established body of literature links low risk perception during pregnancy to reduced facility-seeking behavior [17,18]; this is also consistent with patterns like women with more previous births seeking institutional delivery less (also seen in our study) [19]. When CHW home visits identify no risks, they may inadvertently reinforce the “healthiness” of a pregnancy [20]. This phenomenon could also be more pronounced in a context like South Sudan where health facility access is severely constrained [21]. This could also explain the lack of negative association with institutional delivery in Somalia; Somalia had higher baseline institutional delivery rate than South Sudan, which could be suggesting lower access barriers and possibly less entrenched cultural traditions around childbirth. Given the positive association of the program on select essential newborn care behaviors, we should not conclude that such programs undermine facility-based service utilization, but rather emphasize the importance of concurrent investment in systems and quality strengthening at health facilities as well as strengthen communication strategies that actively promote facility-based care.

### Drivers of the attenuation of positive associations

Notably, almost all key health service indicators collected in our study statistically significantly increased between the baseline and endline surveys, showing positive temporal change across the 1.5-year (South Sudan) and two-year (Somalia) span in which the program was implemented. However, the association between CBMNC exposure and outcomes had mixed results, leaving us with lack of clarity on what is then driving the change. Broader temporal changes affecting both exposed and unexposed women may have contributed to the limited differences observed between CBMNC exposure groups, although this hypothesis cannot be tested directly using the present study design. For example, new or improved health facilities were reported in the programme areas, but their contribution to newborn care practices remains uncertain, given documented constraints in MNH service quality and limited investment in MNH care in both contexts [22]. Furthermore, proportion of women receiving facility-based antenatal care did not statistically change in South Sudan between baseline and endline, suggesting that exposure to facility-based services did not increase in the study population. One hypothesis is social networks; women not enrolled in the program may still have benefited from information spillover through social networks [23–25], and could potentially explain high association between CBMNC and commodity-based behaviors, since information can diffuse through social networks, attenuating the association, whereas physical commodities cannot.

### Increasing salience and desirability of counseling

We observed increase in exposure to counseling as well as uptake of select evidence-based newborn care behaviors, including those that may go against tradition and cultural norms. Existing literature highlights the importance of engaging the holistic environment around the individual, given that these behaviors are embedded in social and cultural norms, family and community influences, and provider attitudes [26]. In both programs, many mechanisms for engaging these “stakeholders” around the woman were executed, which could be one driver behind the increase.

Despite the improvement in behaviors, there are still missed opportunities hinted by both quantitative and qualitative data, the latter presented elsewhere; quantitatively, there are still behaviors like thermal care and 90-day IFA uptake that have notable gaps and qualitatively, the higher perceived value by both demand and supply side of commodity-based interventions versus counseling-based interventions is salient [27]. One study examining the Indian community health workforce (Ashas) observed that Ashas default to behavior change decision-making and persuasion models that do not match with reasons behind why their clients refuse uptake; insufficient or inaccurate understanding of the medical benefits of these services and the dynamics of the social situation were two major reasons behind refusal to uptake, with Ashas defaulting to addressing the former but not the latter [28]. By better equipping CHWs with data on common refusals for uptake and skills for behavior change, their roles could become more effective. We also observed in our data behaviors where the sociocultural origin of the promoted behavior is unclear, like the application on the umbilical cord of tetracycline in Somalia or oil and ash in South Sudan. Further exploration of how those behavioral pathways can be harnessed to promote clinically correct behaviors will be valuable.

### Other community-based interventions

Due to non-permissive policies, there were several evidence-based interventions that were not included. Investment in advocacy and evidence generation to create a more enabling policy and systems environment for community-based service delivery have demonstrated positive outcomes [29,30], and will be invaluable in contexts with high maternal and neonatal mortality. However, inclusion of more services is not necessarily better. Interventions included in the programs described here were selected partially by a statistical modeling approach that attempted to optimize the interventions selected for inclusion in relation to workload, financial cost, training needs, efficacy or effectiveness of the interventions [13]. The statistical modeling, however, was computationally taxing, not to mention the subjectivity of data inputs. There may be more opportunities with artificial intelligence to execute similar analytical procedures more nimbly.

### Limitations

The study was not designed as a quasi-experimental or experimental design, therefore, the observed baseline-endline differences and associations with CBMNC exposure cannot be interpreted as causal. Nevertheless, a key strength was the use of adjusted regression models that accounted for potential confounders, including GPS-based distance to health facilities and selected indicators of seasonal service disruption, thereby reducing the influence of observed confounding. Also, given that enrollment or visit intensity is not randomly assigned and may be driven by some characteristics, the controls for covariates may still leave residual confounding. The data were self-reported and subject to maximum 12-month recall, which could subject the data to recall and social desirability biases; we attempted to mitigate the latter by having data collection conducted by an agency that was not the implementing organization. While we present the data from Somalia and South Sudan in our paper, the two programs differed in multiple ways. The data should be interpreted with those differences in mind. While the two locations represent common rural contexts, the data represent a small geographic area in relation to the whole country.

## Conclusion

CBMNC programs delivered by low-literate CHWs demonstrated high coverage and were associated with meaningful improvements in essential newborn care practices and MNH commodity uptake in rural Somalia and South Sudan. Results were not uniform across settings; in South Sudan, higher antenatal CHW contact was associated with lower odds of institutional delivery, underscoring that community health interventions do not operate in isolation and that parallel investment in facility strengthening and consistent messaging on the complementary roles of community and facility-based care remains essential.

## Data Availability

Data are available at the following sites: https://doi.org/10.6084/m9.figshare.32831078 https://doi.org/10.6084/m9.figshare.32831042

## Acknowledgments

We acknowledge the program implementation teams in both countries (Ahmed Abdi, Hassan Aden Abdi, Inna Caroline, Rachel Douglas, Abdiwahab Maalim, Maryan Miris, Lilian Ndinda, Lual Mayom Nyuany), the Aweil East County Health Department, and the state and national/federal Ministries of Health in South Sudan and Somalia.

