## Appendix File 1 for "Changes in essential newborn care practice and maternal and newborn health commodity uptake following implementation of a community-based maternal and newborn care model in South Sudan and Somalia: pre-post study"

**Detailed description of the community-based maternal and newborn care (CBMNC) intervention packages in Somalia and South Sudan**

| Characteristic | South Sudan | Somalia |
|---|---|---|
| **Overall setting** | | |
| **Implementing partners and governance** | • Delivered by the IRC in close collaboration with the Boma Health Initiative (BHI) and the Reproductive Health Directorate, Ministry of Health, South Sudan.<br>• Intervention package selected through a consultative Constrained Optimisation Workshop held in Juba | • Delivered by the IRC in close collaboration with the Galmudug State and Somalia federal Ministries of Health.<br>• Intervention package selected through a consultative Constrained Optimisation Workshop held in Mogadishu |
| **Community health platform** | • Boma Health Initiative (BHI), the official national structure for community-level service delivery in South Sudan.<br>• Operates under the County Health Department (CHD) within the national Health Sector Transformation Plan. | • Project-recruited female CHW model<br>• CHWs recruited with lower literacy requirements than the national *Marwo Caafimaad* Female Health Worker programme to widen the eligible pool in the target villages. |
| **Setting** | Four *bomas* in Aweil East County, Northern Bahr el Ghazal State | Seven rural villages in Dhusamareb district, Galmudug State, located 20–65 km from the district town |
| **Target population** | Pregnant women residing in the catchment area | Pregnant women residing in the catchment area |
| **Community Health Worker (CHW) cadre** | | |
| **CHW cadre delivering the service** | Boma Health Workers (BHW), the national CHW cadre operating under the Boma Health Initiative. | Female CHWs recruited locally for the project, with lower literacy requirements than the national *Marwo Caafimaad* programme. |

| | | |
|---|---|---|
| **Criteria for CHW selection** | Existing BHWs already operating within the community health system under the CHD and the national Health Sector Transformation Project, with good community standing and an established local social network | Recruited from within the target communities to ensure local acceptability and familiarity with households |
| **Number of CHWs deployed** | 32 BHWs | 34 CHWs |
| **Expected ratio of households served per CHW** | Initially set at 1:50 in line with national policy, but household coverage increased following a policy change immediately before rollout (likely closer to 1:125). | 1:50 |
| **CHW training** | • All BHWs had previously completed the Ministry of Health-mandated two-week BHW training.<br>• For the project, BHWs received a three-day refresher of the BHI Safe Motherhood module followed by a ten-day face-to-face training on the project interventions.<br>• Content drew on BHI national guidelines, WHO community health guidelines, the Essential Newborn Care series and the newborn field guide for humanitarian settings, with video support from the Global Health Media Project.<br>• Competency assessed through a structured pre- and post-test. | • Each CHW provided with a project handbook detailing the required information for every package, used both during training and as an ongoing reference and refresher<br>• Training covered the Federal Ministry of Health community health curriculum, the CBMNC intervention package, data and reporting tools, commodity management, safeguarding, communication, and referral processes. |

| | | |
|---|---|---|
| **Intervention package (overview)** | Six interventions:<br><br>• Prevention of anaemia in pregnancy (dietary counselling and provision of iron-folic acid up to 90 days)<br>• Malaria prevention (distribution of Intermittent Preventive Treatment in pregnancy using Sulfadoxine-Pyrimethamine, or IPTp-SP, and promotion of Long-Lasting Insecticide-treated Nets use).<br>• Advance distribution of oral misoprostol for prevention of postpartum haemorrhage.<br>• Counseling on early initiation of and exclusive breastfeeding<br>• Clean cord care (distribution of chlorhexidine)<br>• Counseling on newborn thermal care (skin-to-skin contact)<br><br>Other general messaging on danger sign identification, birth preparation, and promotion of facility-based services. | Seven interventions:<br><br>• Promotion of basic sanitation and safe drinking water (counseling on handwashing with soap and safe water and sanitation, provision of Aquatabs and soap).<br>• Prevention of anaemia in pregnancy (dietary counselling and provision of iron-folic acid up to 90 days)<br>• Malaria prevention (counseling on bednet use and IPTp-SP and distribution of Long-Lasting Insecticide-treated Nets)<br>• Counseling on early initiation of and exclusive breastfeeding<br>• Counseling on dry cord care<br>• Counseling on newborn thermal care (skin-to-skin contact)<br><br>Other general messaging on danger sign identification, newborn immunization, postpartum family planning, and birth preparation and promotion of facility-based services. |
| **Details of commodities provided** | • Iron-folate tablets: one 30 mg tablet daily, supplied as a 30-tablet monthly refill.<br>• IPTp-SP: at least three doses from the second trimester (not before week 13), at monthly intervals up to delivery, given as directly observed therapy.<br>• Oral misoprostol: 600 mcg (3 x 200 mcg) distributed at the Month 8 visit for home use | • Iron-folate tablets: one 30 mg tablet daily, supplied as one 30-tablet monthly refill, followed by a 60-tablet refill.<br>• Long-Lasting Insecticide-treated Nets: at least one per pregnant woman, distributed at the earliest visit.<br>• Water purification tablets (Aquatabs): 10 per woman per month. |

| | | |
|---|---|---|
| | after delivery where birth occurs at home; unused tablets returned at the postnatal visit.<br>• 7.1% chlorhexidine digluconate gel for cord care, distributed at Month 8 visit.<br><br>Note: Use of Long-Lasting Insecticide-treated Nets is promoted, but nets are distributed through the national net programme rather than by BHWs. | • Soap: 200 g per woman per month.<br><br>• Note: no community IPTp-SP, misoprostol or chlorhexidine gel distributed; IPTp-SP is provided only at facilities and is restricted in high-endemic districts. |
| **Home visit schedule and frequency** | • Target of nine visits per woman: six during the antenatal period and three in the postnatal period, with first contact ideally in the first trimester.<br>• Postnatal visits timed at 24 hours, day 3 and day 7 after birth. | • Target of nine visits per woman: six during the antenatal period and three in the postnatal period, with first contact ideally in the first trimester.<br>• Postnatal visits timed at 24 hours, day 7 and day 14 after birth.<br>• |
| **Maternal danger signs assessed** | • Vaginal bleeding.<br>• Convulsions or fits.<br>• Severe headache with blurred vision.<br>• Fever with inability to get out of bed.<br>• Severe abdominal pain; fast or difficult breathing.<br>• Swelling of the face, fingers and legs.<br>• Thoughts of extreme sadness or of harming the baby. | • Vaginal bleeding.<br>• Convulsions or fits.<br>• Severe headache with blurred vision.<br>• Fever with inability to get out of bed; feeling generally unwell.<br>• Severe abdominal pain; fast or difficult breathing.<br>• Swelling of the face, fingers and legs.<br>• Thoughts of extreme sadness or of harming the baby. |

| | | |
|---|---|---|
| **Newborn danger signs assessed** | • Poor feeding; lethargy; convulsions.<br>• Low body temperature or fever<br>• Chest indrawing; fast breathing (more than 60 breaths per minute)<br>• Signs of an unhealthy cord | • Poor feeding; lethargy; convulsions<br>• Low body temperature or fever<br>• Chest indrawing; fast breathing (more than 60 breaths per minute)<br>• Signs of an unhealthy cord |
| **Referral mechanism** | • BHWs refer to the nearest health facility for maternal or newborn danger signs<br>• Referrals documented in a referral form booklet<br>• Monthly airtime provided to enable ambulance calls | • CHWs refer to the nearest health facility for maternal or newborn danger signs<br>• Referrals documented in a referral form booklet, including newborn referrals<br>• Monthly airtime provided to enable ambulance calls. |
| **Other** | | |
| **Supervision mechanism** | • Quarterly competency assessments were conducted by Supervisors and Safe Motherhood Promoters (SMPs) for all BHWs<br>• Monthly performance review meetings were held with BHWs, Supervisors, and SMPs, focusing on achievements and challenges faced by BHWs<br>• Quarterly joint supportive supervision was conducted with the State Ministry of Health and County Health Department at CBMNC sites. This focused on assessing the skills and knowledge of BHWs in service delivery and identifying operational challenges requiring resolution<br>• Periodic spot checks were conducted by the project team in the field using a supervision checklis | • Quarterly supportive supervision and competency assessments for each CHW<br>• Monthly meetings between CHWs and the program staff<br>• Whatsapp communications in between meetings |
| **Job aids and tools** | • Pictorial job aids and IEC materials adapted for low literacy. | • Pictorial job aids and IEC materials adapted for low literacy. |

| | | |
|---|---|---|
| **provided to CHWs** | • Household registers, referral form booklets<br>• Pregnancy-mapping tool for initial identification of eligible participants at program start | • Project handbook covering all seven intervention packages, used for training and reference.<br>• Household registers, referral form booklets, weekly/monthly data capture tools<br>• Pregnancy-mapping tool for initial identification of eligible participants at program start |
| **Supply chain management** | • Supplies forecast and procured by IRC staff and stored at the IRC warehouse or health facility.<br>• Stock tracked by a program officer using supply tracking data from BHWs with support from the country pharmacist<br>• Kits packed per BHW and replenished monthly | • Supplies forecast and procured by IRC staff and stored at the IRC warehouse or health facility.<br>• Stock tracked by a program officer using supply tracking data from CHWs with support from the country pharmacist<br>• Kits packed per CHW and replenished monthly |
