## Appendix File 2 for "Changes in essential newborn care practice and maternal and newborn health commodity uptake following implementation of a community-based maternal and newborn care model in South Sudan and Somalia: pre-post study"

**Appendix File 2: Creation of household wealth index**

Household socioeconomic status was measured using an asset-based wealth index constructed separately for each country (Somalia and South Sudan) and survey round (baseline and endline) using principal components analysis (PCA). The index was based on binary indicators of household ownership of durable assets and housing characteristics collected in the household survey.

Candidate variables included ownership of household assets (e.g., radio, mobile phone, smartphone, watch, bed, Somali stool, sitting cushion, electricity), housing construction materials (floor and wall materials), and access to improved drinking water. An improved drinking water indicator was created according to WHO/UNICEF Joint Monitoring Programme (JMP) classifications by combining improved water sources into a single binary variable and distinguishing them from unimproved sources. Variables with no or negligible variation (e.g., ownership prevalence close to 0% or 100%), variables with substantial structural missingness (e.g., internet access collected only among mobile phone owners), and redundant complementary indicators (e.g., both natural and finished housing materials) were excluded from the PCA to maximize discriminatory power and avoid collinearity.

Because the distribution of household assets differed substantially across countries and survey rounds, the final set of variables included in the PCA was tailored to each dataset while following a consistent selection strategy. In Somalia, the wealth index incorporated household assets (electricity, watch, mobile phone, bed, Somali stool, sitting cushion), improved drinking water, and finished floor and wall materials. In South Sudan, where considerably fewer household assets demonstrated sufficient variability, the wealth index was based on radio ownership, mobile phone ownership, smartphone ownership, watch ownership (baseline only), improved drinking water, and finished wall materials (endline only).

This approach is consistent with the asset-based wealth indices commonly used in Demographic and Health Surveys (DHS), although the specific assets included were adapted to those available in each survey.

PCA was performed using the correlation matrix of the selected binary variables, and the first principal component was retained as the continuous household wealth score. Higher scores indicate greater household socioeconomic status. Households were classified into two wealth groups (lower and higher wealth) based on the median of the PCA score within each country and survey round. In South Sudan only, because the limited number of assets resulted in tied PCA scores at the median, ties were broken using a reproducible random ordering with a fixed random seed to ensure equal-sized wealth groups.

***Figure A- 1: Distribution of household wealth index scores by country and survey round***

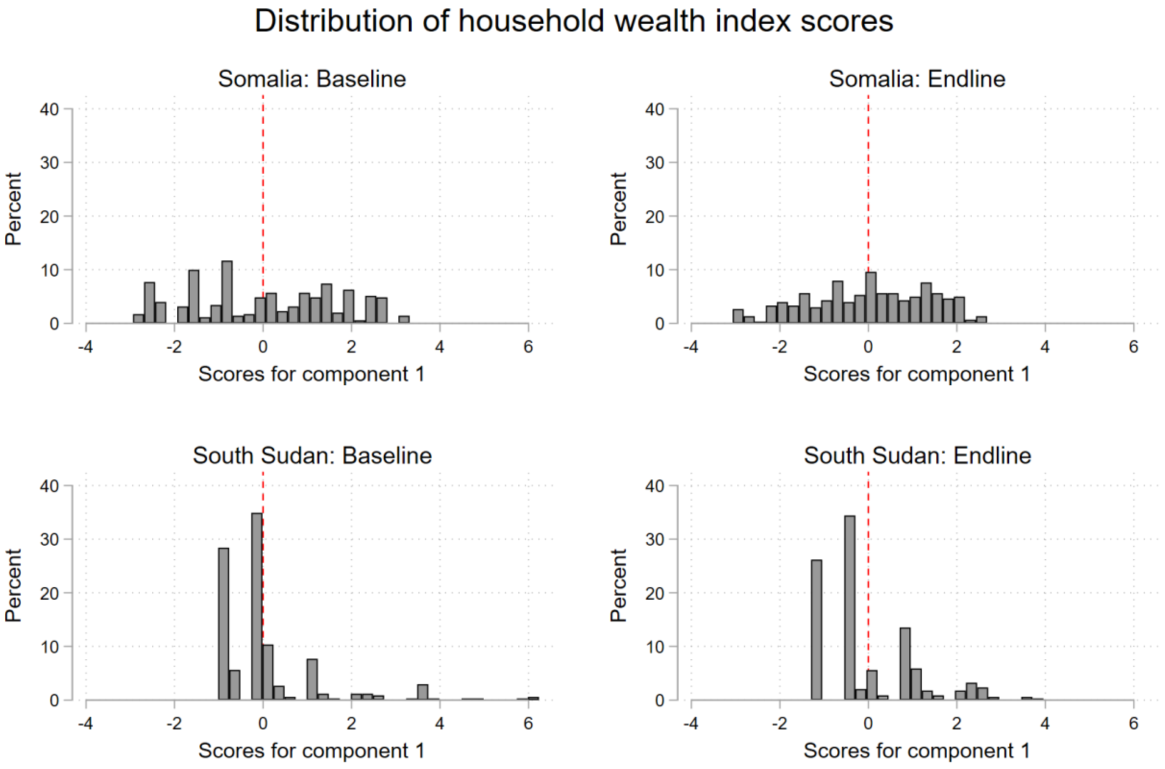
